# Role of COVID-19 vaccinal status in admitted children during OMICRON variant circulation in Rio de Janeiro, city- Preliminary report

**DOI:** 10.1101/2022.02.10.22270817

**Authors:** AR Araujo da Silva, BRR de Carvalho, MM Esteves, CH Teixeira, CV Souza

## Abstract

**Objective:** To evaluate COVID-19 vaccination status in admitted children during the OMICRON variant circulation.

**Design:** Observational retrospective study. Patients with confirmed COVID-19 were compared in two different periods: 2020-2021 when full COVID-19 vaccine were not completed for adolescents between 12 and 18 years and 2022, when Pfizer-BioNTech vaccine full scheme were completed for children older 12 years

**Setting:** Two paediatric hospitals in Rio de Janeiro, city

**Patients:** Children aged <18 years with confirmed COVID-19.

**Intervention:** None

**Main outcome:** Vaccination status for COVID-19 at the admission.

**Results:** Three-hundred patients were admitted with confirmed COVID-19, being 240 in 2020-2021, and 60 in 2022. The distribution of patients according to the age-groups were: 0-2 years (33.3% in 2020-2021 and 53.4% in 2022), 2-5 years (21.7% in 2020-2021 and 10% in 2022), 5-11 years (29.2% in 2020-2021 and 28.3% in 2022) and 12-18 years (15.8% in 2020-2021 and 8.3% in 2022) (p= 0.015). The median of lenght of stay was 6 days in 2020-2021 and 5 days in 2022 (p=0.036). We verified 6 deaths and in the first period of analysis and 1 death in the second one (p=0.777). Of 60 children admitted in 2022, fifty-eigth (96.7%) not received the full scheme of COVID-19 vaccine available

**Conclusions:** We verified in a “real world condition” ability of Pfizer-BioNTech vaccine in avoid hospitalization in children older than 12 years

## Introduction

COVID-19 pandemic is still in course by January 2022, after more than two years of the initial description of SARS-COV-2 ^1^. Since then, at least five variants of concern spread around the world, being the B.1.1.529 variant (OMICRON) the most prevalent in most countries, including Brazil, since the first report in 24 November 2021, in South Africa ^2^.

According to the Brazilian Health Ministery, until the 48^th^ epidemiological week (December 4, 2021) 1,422 children died in consequence of acute respiratory syndrome distress due to COVID-19, which represent 0.38% of total of deaths (372,954) ^3^. Despite of relative low relative number of deaths, the absolute number of children deaths due to COVID-19 was 8.5 times higher than all other respiratory virus deaths together causing the same syndrome ^3^.

One important strategy to minimize COVID-19 hospitalization and deaths is COVID vaccination, recently evaluated in adolescents and children older than 5 years ^4,5^. The COVID-19 campaing for children between 12 and 17 years-old in Brazil, started in August 2021 with BNT162b2 (Pfizer-BioNTech) vaccine administered in two doses with 12 weeks apart. In 17 January 2022, the COVID-19 vaccination for children between 5 and 11 years old began in Brazil. As effect of COVID-19 in children need to be studied, in a “real world settings” regarding distribution age of admitted children and outcomes, our aim is to evaluate COVID-19 vaccination status in admitted children during the OMICRON variant season.

## Methods

We conducted an observational retrospecty study in children from 0 to 18 years-old admitted in two pediatric hospitals of Rio de janeiro city, Brazil. The hospitals receive patients from his own emergency departments or refered to hospitalization (in wards or intensive care units) of other units. The unit 1 and 2 has 135 and 45 bed-capacity, respectively. Children admitted more than 24h between 1^st^ January 2020 and 10^th^ February 2022, in any ward, were included if filled the WHO case definitions ^6^. Children admitted but transfered for other hospitals were excluded.

In Rio de Janeiro city, COVID-19 vaccination of children between 12 and 17 years was completed (two-dose) only in mid December 2021. The children between 5 to 11 years-old started to received COVID-19 vaccine in 17th January, 2022. The BNT162b2 (Pfizer-BioNTech) vaccine and CoronaVac are until the moment the vaccines approved in Brazil for administration in children of 5 to 18 years-old age, and for children of 6 to 18 years-old age, respectively. Variant identification of positive cases was not conducted.

Distribution age of confirmed cases were compared with the period before of COVID-19 vaccines availabilty for children between 12 and 18 years (2020 and 2021). Considering that full protection for children between 12 and 18 years were obtained only in 30th December 2021, we admit that all children with confirmed COVID-19 were not fully protected against SARS-COV-2 in 2020 and 2021 years.

A descriptive analyses of age, gender, lengh of stay, outcomes and vaccinal status was performed using Microsoft Excel. When appropriate, we used Chi square test for categorical variables and Mann–Whitney U test for continuous variables. A value of p less than .05 were considered as statistical significant.

The study was submitted and approved by Ethics Committee of Faculty of Medicine (Universidade Federal Fluminense) and Prontobaby Group, under number 4.100.232 dated from June 20, 2020. A consent statement for the use of data from the patients or their parents or guardians were required for each children included.

## Results

Three hundred patients were admitted with confirmed COVID-19, being 240 in 2020 and 2021, and 60 in 2022 (until 10^th^ February). The demographic data of confirmed patients,lenght of stay and outcomes are shown in table 1

**Table 1-.** Demographic data of admitted cases in two pediatric hospitals (Rio de Janeiro city, 1^st^ January 2022-15^th^ February 2022)

|  | 2020/2021<br>N=240 | 2022<br>N=60 | P value |
| --- | --- | --- | --- |
| **Gender** |  |  |  |
| -Female | 104 (43.3) | 30 (50) | 0.352 |
| -Male | 136 ( 56.7) | 30 (50) |  |
| **Age** |  |  |  |
| 0-2 years | 80 (33.3) | 32 (53.4) | **0.015** |
| 2-5 YEARS | 52 (21.7) | 6 (10) |  |
| 5-11 years | 70 (29.2) | 17 (28,3) |  |
| 12-18 years | 38 ( 15.8) | 5 (8.3) |  |
| **Length of stay ( MEDIAN in days)** | 6 (1-164) | 5 (1-19) | **0.036** |
| **Outcome after 7 days\*** |  |  |  |
| - Discharged | 234 (97.5) | 53 (88.3) | 0.777 |
| - Death | 6 (2.5) | 1 ( 1.7) |  |

\*6 patients still hospitalized in 2022

Of five patients older than 12 years, that were admitted in 2022, 2 received two-dose, 2 received one-dose and 1 patient recieved zero-dose. All patients admitted in 2020 and 2021 year were not fully immunizated.

## Discussion

The control of COVID-19 in children includes the capacity of vaccines to minimize acquisition of disease and reduce hospitalization and deaths. In our study, we verified a clear pattern change pattern of confirmed cases admitted in 2022 year, according to the distribution age (after fully vaccination of children older than 12 years old age). Of 60 children admitted in 2022, only two of them (3.3%) received the full scheme of COVID-19 vaccine available. Only in middle of January 2022, COVID-19 vaccines started to be administrated in children between 5 and 12 years.

The pattern of lenght of stay was also modified in 2022, being statistically shorter than verified in the two previous years. Although this is an important marker, a longer period of analysis could confirm this finding, considering that six children still hospitalized at the end of this report.

In the first six weeks of 2022, Rio de Janeiro city and Brazil experienced a higher circulation of OMICRON variant, causing increase of number cases in all ages, including children. We verified that mortality due to COVID-19 was similar in the two periods studied, but the true impact of COVID-19 vaccines in reducing mortality in children needs to be accessed in prospective studies, evaluating patients after vaccination for different children ages.

In conclusion, we verified in a “real world condition” ability of Pfizer-BioNTech vaccine in avoid hospitalization in children older than 12 years

## Data Availability

All data produced in the present study are available upon reasonable request to the authors

## References

1. Zhu N, Zhang D, Wang W, Li X, Yang B, Song J, et al. A Novel Coronavirus from Patients with Pneumonia in China, 2019. N Engl J Med. 2020 Feb 20;382(8):727–733.

2. Classification of Omicron (B.1.1.529): SARS-CoV-2 Variant of Concern. World Health Organization. Available at: https://www.who.int/news/item/26-11-2021-classification-of-omicron-(b.1.1.529)-sars-cov-2-variant-of-concern. Accessed on January 16, 2022.

3. BOLETIM EPIDEMIOLÓGICO ESPECIAL-92. Doença pelo Novo Coronavírus – COVID-19. Secretaria de Vigilância em Saúde, MINISTÉRIO DA SAÚDE-Brazil. Available at: https://www.gov.br/saude/pt-br/centrais-de-conteudo/publicacoes/boletins/boletins-epidemiologicos/covid-19. Accessed on January 16, 2022.

4. Frenck RW Jr, Klein NP, Kitchin N, Gurtman A, Absalon J, Lockhart S, Perez JL, Walter EB, Senders S, Bailey R, Swanson KA, Ma H, Xu X, Koury K, Kalina WV, Cooper D, Jennings T, Brandon DM, Thomas SJ, Türeci Ö, Tresnan DB, Mather S, Dormitzer PR, Şahin U, Jansen KU, Gruber WC; C4591001 Clinical Trial Group. Safety, Immunogenicity, and Efficacy of the BNT162b2 Covid-19 Vaccine in Adolescents. N Engl J Med. 2021 Jul 15;385(3):239–250.

5. Walter EB, Talaat KR, Sabharwal C, Gurtman A, Lockhart S, Paulsen GC, Barnett ED, Muñoz FM, Maldonado Y, Pahud BA, Domachowske JB, Simões EAF, Sarwar UN, Kitchin N, Cunliffe L, Rojo P, Kuchar E, Rämet M, Munjal I, Perez JL, Frenck RW Jr, Lagkadinou E, Swanson KA, Ma H, Xu X, Koury K, Mather S, Belanger TJ, Cooper D, Türeci Ö, Dormitzer PR, Şahin U, Jansen KU, Gruber WC; C4591007 Clinical Trial Group. Evaluation of the BNT162b2 Covid-19 Vaccine in Children 5 to 11 Years of Age. N Engl J Med. 2022 Jan 6;386(1):35–46.

6. WHO COVID-19: Case Definitions. World Health Organization. Available at: https://apps.who.int/iris/rest/bitstreams/1322790/retrieve. Accessed on January 16, 2022

